# Impact of a Community-Based Positive Deviance Hearth Intervention on Feeding Practices Among Malnourished Children Aged 6–59 Months in Bomet County, Kenya

**DOI:** 10.64898/2026.04.18.26351171

**Authors:** Gladys C. Koskei, Simon Karanja, Zipporah W. Ndungu, Calvince O. Anino

## Abstract

Child undernutrition remains a major public health challenge in Kenya. Suboptimal feeding practices contribute significantly to persistent underweight and stunting. This study evaluated the effect of a community-based Positive Deviance Hearth (PDH) intervention on feeding practices among children aged 6–59 months in Sub County within a County of study. The study adopted a two-group pretest-posttest randomized experimental study design conducted for six months period, among 84 caregiver-child pairs in intervention and control groups. A multi-stage sampling was employed to identify study settings and participants. Structured and pretested questionnaires, 24-hour food recall questionnaires and meal diversity questionnaires were used for data collection at pre-intervention and post-intervention periods. Data was analyzed using R software *v*.*4*.*5*.*2*. The differences between intervention and control groups at baseline and endline were assessed using difference-in-difference analysis, relevantly summarized using adjusted DID estimates, 95% confidence intervals and p-values, with p<0.05 considered significant. The PDH intervention significantly improved feeding practices among children 6–59 months. Meal frequency increased for 9–23 months (DiD = +1.4; 95% CI: 1.2–1.7; p = 0.034) and ≥24 months (DiD = +1.2; 95% CI: 1.1–1.5; p = 0.017), and dietary diversity rose (DiD = +1.3; 95% CI: 1.1–1.9; p < 0.001). Nutrient-dense food consumption improved, including legumes (DiD = +32.6%; p < 0.001) and animal-source foods (DiD = +35.4%; p < 0.001). Energy and protein intake increased across all age groups (p < 0.05), and micronutrients—iron, vitamin A, vitamin C—also rose significantly (p < 0.05). The PDH intervention substantially improved caregiver feeding practices, increased dietary diversity, and enhanced macro- and micronutrient intake, demonstrating its effectiveness as a scalable, community-driven strategy for sustainably improving child nutrition in high-burden settings.

## INTRODUCTION

Child undernutrition remains a major global public health problem, disproportionately affecting low- and middle-income countries. Undernutrition contributes to nearly a half of all deaths among under five children, especially through increased susceptibility to infectious diseases (1,2). Malnutrition significantly impacts child survival, growth and development, particularly through impaired cognitive development, declined educational performance, and economic productivity across the life course (3). In 2023, approximately 150 million children under five years were stunted, 44 million were wasted, and 38 million were overweight globally (1,4). Despite global commitments under United Nations Sustainable Development Goal 2 (Zero Hunger) and Goal 3 (Good Health and Well-being), progress in childhood malnutrition reduction remain uneven, especially in sub-Saharan Africa (5). This has been attributed to suboptimal infant and young child feeding (IYCF) practices, food insecurity, structural poverty and limited access to quality health services. The burden of child undernutrition remains a major concern in Kenya, with 18% were stunted, 10% were underweight, and 5% were wasted in 2022 (6). However, these averages obscure higher burdens in rural and food-insecure counties like Bomet county. According to Korir et al.(7) and Mwangangi et al.(8), study area is characterized by poverty and agrarian livelihoods, household food insecurity and cultural feeding norms which contribute to suboptimal child feeding practices, food access and dietary diversity in the region. Persisted malnutrition among under five has been reported despite ongoing public health efforts (6). According to KHIS of 2018-2022, reported persistent malnutrition in the sub-county affecting more than 3,500 under five children, attributed to poor maternal education, cultural beliefs and household food insecurity.

Optimal feeding practices during the first 1,00 days of life is critical and cost-effective approach for malnutrition prevention. According to World Health Organization, exclusive breastfeeding should be done for the first six months, followed by the introduction of safe, adequate and nutritious complementary foods while continuing breastfeeding up to two years and beyond (9). However, adherence to these guidelines remains suboptimal, and translating knowledge into sustainable behaviour change is a persistent challenge. Conventional nutrition education and related interventions such as supplementation and food aid programs have often failed to effectively address child malnutrition (10,11). Similarly, these approaches often do not account local socio-cultural, economic constraints, and intra-household decision-making processes, hence limiting their effectiveness, sustainability, scalability, and cultural acceptability (12). As a result, community-driven, behavior-focused approaches that leverage existing local solutions and lived-realities have drawn significant attention over the past few years.

The Positive Deviance Hearth (PDH) approach is based on the observation that within malnutrition high-risk communities, some individuals or households achieve better health and nutrition outcomes through uncommon but successful behaviors despite facing similar constraints as their peers (13,14) The PDH model identifies locally available, acceptable, affordable and beneficial feeding and caregiving practices from positive deviant caregivers, and disseminates them through structured participatory, community-driven sessions. During these sections, caregivers of malnourished children engage in participatory learning, cooking demonstrations and preparations, and practical feeding activities (15,16). PDH interventions have reported significant improvement in dietary intake, caregiver knowledge and nutrition outcomes among undernourished children (12,15). However, these findings remain varying across PDH studies. Some studies have reported short-term gains that are not sustainable over time and/or without sustained behavioral change, while others outlining challenges related to program’s scalability, long-term impacts, and contextual adaptation especially in regions prone to food insecurity and limited follow-up support (17,18). Similarly, there is limited empirical evidence on impact of PDH interventions on feeding practices, as few PDH studies in Kenya have focused on nutritional outcomes (12,19). This is further influenced by contextual variations in socio-cultural norms, food systems, and health service delivery. Therefore, there is need for controlled study design to evaluate PDH impact, for robust evidence generation for practice and policy. This study assessed the impact of a community-based PDH intervention on feeding practices among malnourished children aged 6–59 months.

## MATERIALS AND METHODS

### Study Design and Settings

The study adopted a two-group pretest-posttest randomized experimental study design conducted for six months period, from March 2025 to September 2025. The study involved intervention and control groups. The design allowed comparison of the two groups while controlling for baseline differences. The study was conducted in at sub-County level in Kenya. The study setting face persists burden malnutrition among under five children, with prevalence of 22% in 2022 (6).

This could be attributed to rural agrarian livelihoods, relatively higher poverty levels, food insecurity due to relance to cash crop (tea) farming and suboptimal IYCF practices (7,8).

### Study Population and Eligibility Criteria

The study involved caregiver-child pairs, where identified moderately malnourished children aged 6-59 months were included. This age group is a critical developmental window during which children become highly susceptible to growth faltering, micronutrient deficiencies, and infections (1,2). Similarly, it’s at this age group when growth is rapid hence highest nutrient requirement. The study included permanent residents of selected the two wards, and caregivers who consented to participate. However, the study excluded children with severe medical conditions or congenital abnormalities requiring specialized care and influencing feeding.

### Sample Size Determination and Sampling

Sample size was determined using Fleiss formula for comparing two proportions, n = {[P_1_(1−P_1_) + P_2_(1−P_2_)] × f(α, β}/(P_1_ – P_2_)^2^, where *f*(*α,β*) = 10.5 at 95% confidence and 80% power (20). Using a baseline prevalence of underweight in the study area of *P*_1_= 0.10 (6) and an anticipated post-intervention prevalence of *P*_2_ = 0.012, the calculated sample size was *n* = 139. Allowing for 20% attrition increased the total sample to *n* = 167, which was equally allocated between study arms, yielding 84 participants per group. A multi-stage sampling approach was used, where the sub-County was purposively selected due to high malnutrition burden, two wards using simple random sampling, and participants using simple random sampling technique from list generated after community case identification through anthropometric measurements and categorization. Active community case identification was conducted for three weeks by community health promoters (CHPs), community nurses and principal investigators.

### Description of the Intervention

The PDH intervention was implemented as a structured, community-based behavior change package grounded in the positive deviance approach. A preliminary formative inquiry identified context-specific, feasible feeding and caregiving practices from positive deviant households, those with well-nourished children despite comparable socio-economic constraints. These practices informed a 12-day intensive hearth model delivered in small caregiver groups and facilitated by trained community health volunteers (CHVs) and community nurses from 9^th^ to 21^st^ March 2025. Each session integrated participatory learning, live food demonstrations using locally available ingredients, supervised child feeding, and practical coaching on age-appropriate IYCF practices, food preparation, hygiene, and responsive caregiving. Emphasis was placed on experiential learning, peer reinforcement, and repeated practice to enhance skill acquisition and behavioral adoption. Hearth education sessions also involved use of educational videos, pictures and relevant audio-visuals which were projected during these sessions. This approach was chosen since none of participants had visual nor hearing impairment, and it was more effective. Following the group sessions, caregivers received scheduled home visits to reinforce adherence, address contextual barriers, and monitor translation of learned practices into routine household behavior, thereby strengthening sustainability of the intervention effects. Follow-up session were conducted for aa period of six months. The control group received conventional or usual community health services. They were not subjected to specific PDH intervention.

### Data Collection Methods

Data were collected using a structured, pretested questionnaire administered through interviewer-led, face-to-face interviews with primary caregivers by trained research assistants. The tool was specifically designed to capture feeding practices, including breastfeeding behaviors, complementary feeding patterns, feeding during child illness, food preparation, feeding frequency, portioning, and responsive feeding techniques. A standardized 24-hour dietary recall was employed to quantify recent intake, from which mean dietary diversity scores and consumption of key food groups (grains, roots and tubers; legumes; milk and dairy products; fish, eggs, meat and other animal-source foods; vitamin A–rich fruits and vegetables; and other fruits and vegetables) were derived. Age-specific estimates of macronutrient intake (energy, protein, and carbohydrates) and selected micronutrients (calcium, iron, zinc, vitamin A, vitamin C, and folate) were computed using standard food composition references. Data were collected at baseline (pre-intervention), on the third month (midline) and sixth month (endline) across all study groups. Baseline assessment was conducted from January to February 2025, midline on June 2025, while on September 2025 endline data collection was conducted.

### Data Quality Assurance

Multi-tiered approach was adopted to ensure data quality. Research assistants, who understood local dialect and experienced in nutrition research, and community health volunteers underwent intensive five days training on the study objectives, ethical participant recruitment and engagement, questionnaire administration, 24-hour dietary recall techniques, portion quantification, and measurement of specific indicators. A pilot study was conducted in a different ward with similar socio-economic and demographic characteristics within the same sub-county involving 20 pairs of caregiver and malnourished child aged under five years. This was done two weeks prior main study. Pretest data demonstrated the Cronbach alpha coefficient of 0.73, considered reliable. Supervisors, led by principal investigator, conducted daily spot checks, observed interviews, and assessed completeness and consistency of responses during data collection fieldwork. Portion sizes were measured using calibrated utensils and standardized measures to minimize risks of measurement errors. Additionally, data were double-entered, cross-validated, and carefully cleaned to minimize potential inconsistencies.

### Data Management and Analysis

Cleaned data were analyzed using R software *v*.*4*.*5*.*2*. Continuous variables, including dietary diversity scores (DDS) and age-specific nutrient intakes, were summarized as means ± standard deviations, while categorical feeding behaviors were expressed as frequencies and percentages. DDS was calculated by summing the number of key food groups consumed in the preceding 24 hours: grains, roots and tubers; legumes and nuts; dairy products; flesh foods; eggs; vitamin A-rich fruits and vegetables; and other fruits and vegetables. Difference-in-difference (DiD) analysis was conducted to evaluate the effect of the PDH Intervention on feeding practices, with multivariate linear DiD models for continuous outcomes and logistic DiD models for binary/categorical feeding behaviors. Each model included study group, time point, and the interaction term, which served as the adjusted DiD estimator with 95% confidence intervals and p-values. All analyses were conducted at a 95% confidence level (p < 0.05).

### Ethical consideration

Ethical approval was obtained from the institutional scientific and ethics review committee (UEAB/ISERC/07/08/2024) and permit from National Commission for Science Technology and Innovation (NACOSTI/P/24/39809). Additionally, permission was sought from county ministry of health and local leadership. Informed consent was obtained from the caregivers/guardians of all the children. The investigators explained the purpose, benefits, duration and study related activities to all participants. Participation was voluntary and participants were informed that they can withdraw from the study without consequences. Confidentiality and privacy as ensured through anonymization of data and private interviews.

## RESULTS

Out of 84 participants included in each arm, only 76 participants in the intervention group and 79 participants in control groups participated to completion of the study. There was no significance difference in socio-demographic characteristics of the participants at baseline and endline between control and intervention groups (p>0.05). The study participants had similar characteristics (Table 1).

**Table 1.**
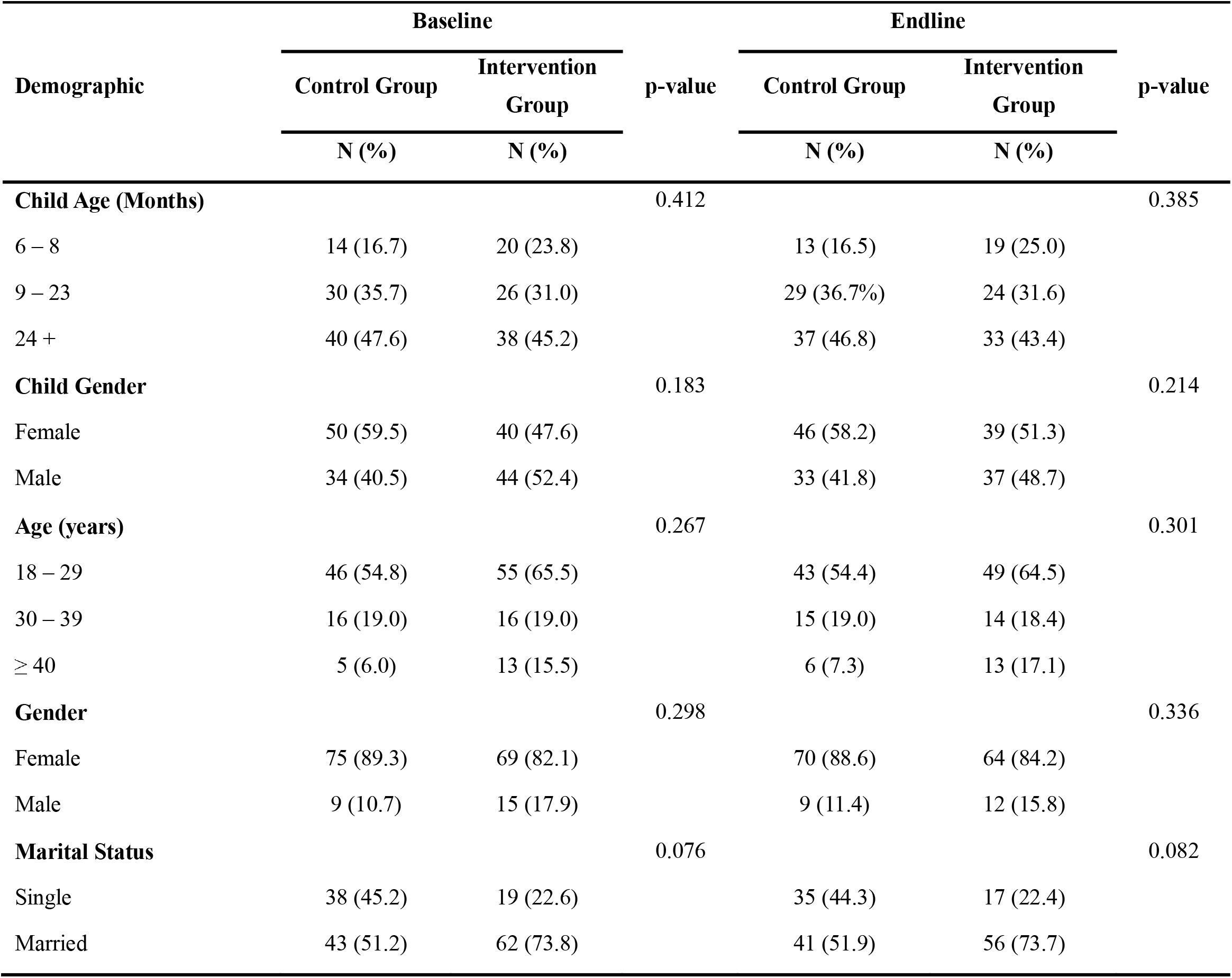
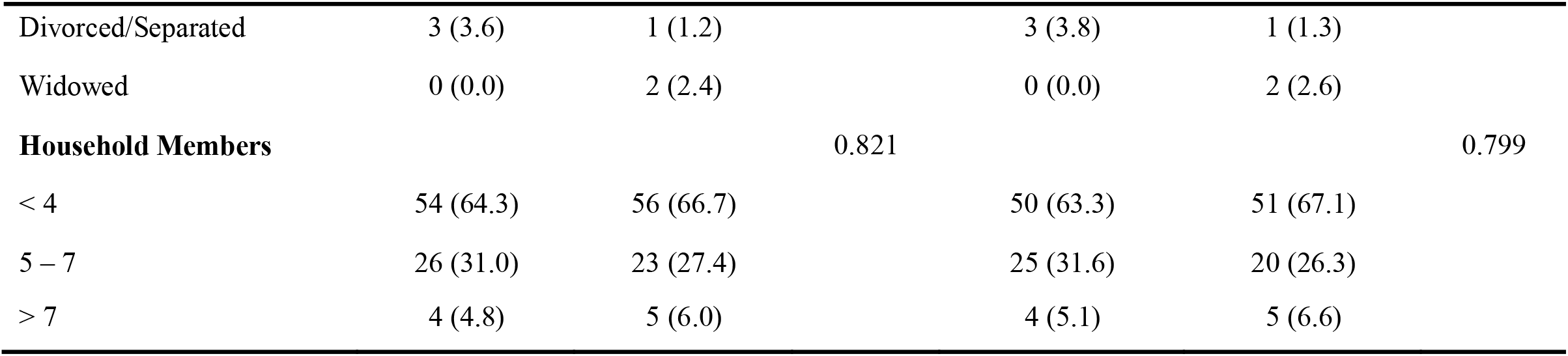
Demographic characteristics of children and care givers at baseline of the study.

### General Feeding Practices

The PDH intervention had significant improvement on feeding practices (Table 2). Compared to control groups, the intervention significantly improved meal frequency among children aged 9-23 months (DID: +1.4; 95% CI: 1.2–1.7; p = 0.034) and those aged ≥24 months (DID: +1.2; 95% CI: 1.1–1.5; p = 0.017) after six months. Similarly, dietary diversity scores improved among intervention group compared to control group (DID = +1.3; 95% CI: 1.1–1.9; p < 0.001). The PDH intervention resulted to significant improvement in appropriate responsive feeding practices among caregivers by 33.9% (95% CI: 24.7–43.1; p < 0.001) and 34.9% increase in preparation of child-specific meals (95% CI: 25.8–44.0; p < 0.001).

**Table 2.**
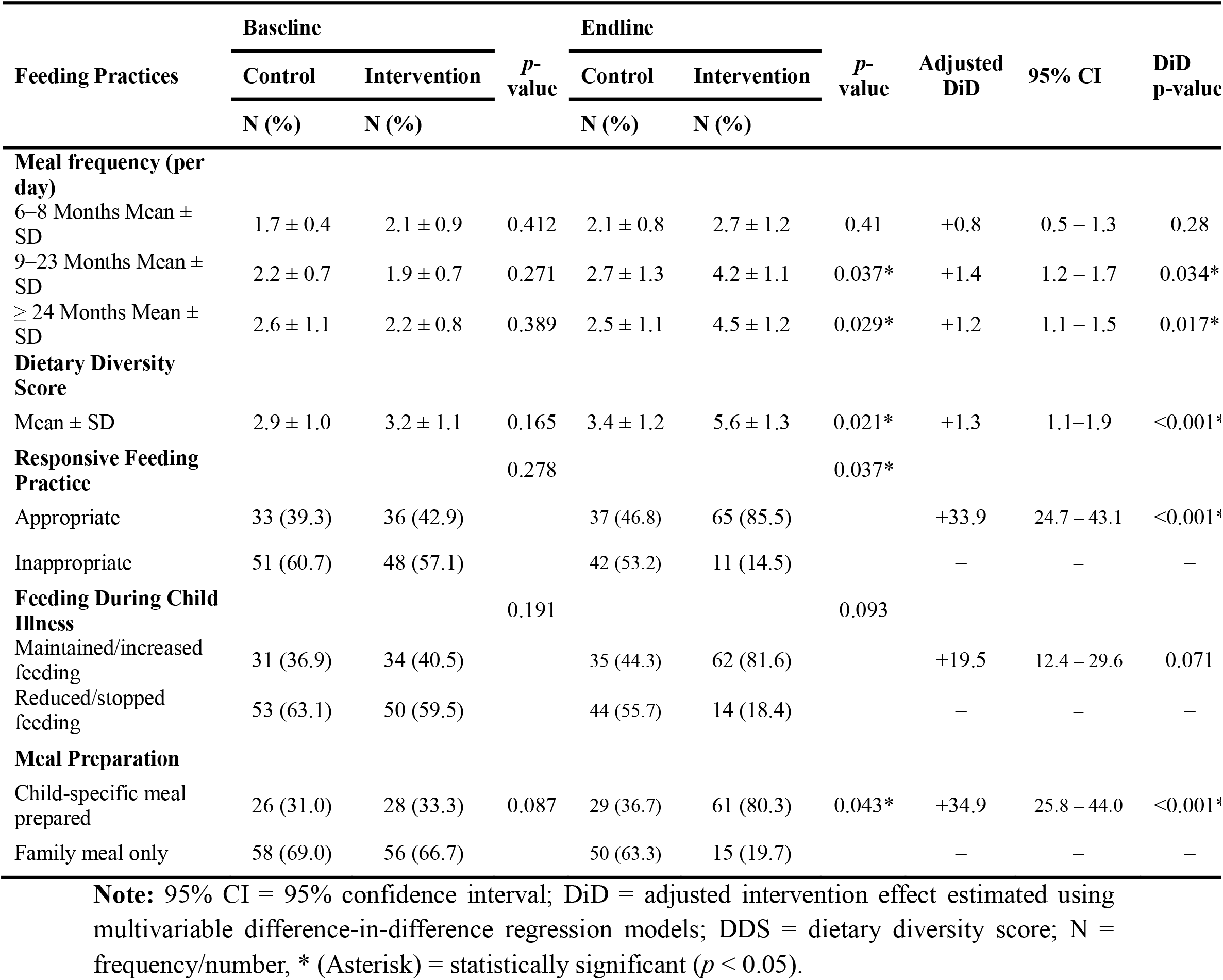
Feeding practices among participants in intervention and control groups.

### Food Group Consumption among participants

The participants heavily relied on grains, roots and tubers (≥90%) and milk and dairy products (71.4 −72.6%) with low intake of legumes, animal-source foods, and fruits and vegetables at baseline (p>0.05). At endline, adjusted difference-in-differences analyses demonstrated that the intervention produced statistically significant improvements in the consumption of nutrient-dense food groups, including legumes (DID: +32.6%, 95% CI: 18.9–44.1; p<0.001), fish/eggs/meat and other animal-source foods (DID: +35.4%, 95% CI: 21.6–47.3; p<0.001), vitamin A–rich fruits and vegetables (DID:+30.1%, 95% CI: 15.7–42.8; p=0.001), and other fruits and vegetables (DID: +28.7%, 95% CI: 14.3–41.0; p=0.002). In contrast, no statistically significant intervention effects were observed for grains, roots and tubers or milk and dairy products (Table 3).

**Table 3.** Difference-in-differences analysis of food group consumption derived from dietary diversity questionnaire among 6-59 children.

| Food Groups | Baseline |  | *p*-value | Endline |  | *p*-value | Adjusted DiD | 95% CI | DiD p-value |
| --- | --- | --- | --- | --- | --- | --- | --- | --- | --- |
|  | Control | Intervention |  | Control | Intervention |  |  |  |  |
|  | N (%) | N (%) |  | N (%) | N (%) |  |  |  |  |
| **Grains, roots and tubers** |  |  | 0.812 |  |  | 0.644 |  |  |  |
| Yes | 78 (92.9) | 77 (91.7) |  | 72 (91.1) | 70 (92.1) |  | +4.5 | −3.2–12.1 | 0.248 |
| No | 6 (7.1) | 7 (8.3) |  | 7 (8.9) | 6 (7.9) |  | – | – | – |
| **Legumes** |  |  | 0.604 |  |  | 0.004* |  |  |  |
| Yes | 29 (34.5) | 31 (36.9) |  | 30 (38.0) | 53 (69.7) |  | +32.6 | 18.9 – 44.1 | <0.001* |
| No | 55 (65.5) | 53 (63.1) |  | 49 (62.0) | 23 (30.3) |  |  |  |  |
| **Milk and dairy products** |  |  | 0.781 |  |  | 0.219 |  |  |  |
| Yes | 61 (72.6) | 60 (71.4) |  | 56 (70.9) | 59 (77.6) |  | +6.8 | −4.6–18.2 | 0.241 |
| No | 23 (27.4) | 24 (28.6) |  | 23 (29.1) | 17 (22.4) |  | – | – | – |
| **Fish/Eggs/Meat and animal products** |  |  | 0.543 |  |  | 0.002* |  |  |  |
| Yes | 18 (21.4) | 20 (23.8) |  | 19 (24.1) | 45 (59.2) |  | +35.4 | 21.6–47.3 | <0.001* |
| No | 66 (78.6) | 64 (76.2) |  | 60 (75.9) | 31 (40.8) |  | – | – | – |
| **Vitamin-A rich fruits and vegetables** |  |  | 0.417 |  |  | 0.006* |  |  |  |
| Yes | 24 (28.6) | 27 (32.1) |  | 26 (32.9) | 47 (61.8) |  | +30.1 | 15.7–42.8 | 0.001* |
| No | 60 (71.4) | 57 (67.9) |  | 53 (67.1) | 29 (38.2) |  | – | – | – |
| **Other fruits or vegetables** |  |  | 0.502 |  |  | 0.008* |  |  |  |
| Yes | 31 (36.9) | 33 (39.3) |  | 32 (40.5) | 53 (69.7) |  | +28.7 | 14.3–41.0 | 0.002* |
| No | 53 (63.1) | 51 (60.7) |  | 47 (59.5) | 23 (30.3) |  | – | – | – |

Note: CI = confidence interval; DiD = difference-in-differences; DDS = dietary diversity score; N = frequency; SD = standard deviation; \* (asterisk)– statistically significant difference at $p < 0.05$ .

### Macronutrient Intake among Under-Five Children

At baseline, there were no significant differences in macronutrient intake between intervention and control groups across all age categories (p > 0.05). However, by endline, significant improvements were observed among children in the intervention group (Table 4). Infants 6– 11 months consumed 619 ± 109.4 kcal compared with 538 ± 101.2 kcal in controls (p = 0.043) and 9.2 ± 1.8 g protein versus 6.3 ± 1.5 g (p = 0.021). Children 12– 36 months in the intervention group had higher energy intake (903 ± 169.6 kcal) than controls (871 ± 157.8 kcal, p = 0.007) and protein intake increased from 7.6 ± 2.5 g to 16.1 ± 3.0 g (p = 0.015). Among 37–59 months, protein intake rose to 18.3 ± 3.0 g from 8.5 ± 2.4 g in intervention group (p = 0.016).6

**Table 4.** Age-specific Macronutrient Intake among Under-Five Children at Baseline and Endline of the Study.

| Age Group<br>(Months) | Nutrient | RDA | Baseline |  | p-value | Endline |  | P-value |
| --- | --- | --- | --- | --- | --- | --- | --- | --- |
|  |  |  | Control | Intervention |  | Control | Intervention |  |
|  |  |  | (Mean ± SD) | (Mean ± SD) |  | (Mean ± SD) | (Mean ± SD) |  |
| 6–11 |  |  |  |  |  |  |  |  |
|  | Energy (kcal) | 710 | 528 ± 98.4 | 533 ± 101.6 | 0.687 | 538 ± 101.2 | 619 ± 109.4 | 0.043* |
|  | Protein (g) | 10 | 5.9 ± 1.4 | 6.1 ± 1.3 | 0.613 | 6.3 ± 1.5 | 9.2 ± 1.8 | 0.021* |
|  | Carbohydrates (g) | 95 | 76.4 ± 13.8 | 75.9 ± 14.6 | 0.781 | 78.3 ± 14.9 | 93.1 ± 16.2 | 0.089 |
| 12–36 |  |  |  |  |  |  |  |  |
|  | Energy (kcal) | 1300 | 856 ± 148.7 | 841 ± 136.2 | 0.721 | 871 ± 157.8 | 903 ± 169.6 | 0.007* |
|  | Protein (g) | 16 | 7.3 ± 2.1 | 7.5 ± 2.3 | 0.652 | 7.6 ± 2.5 | 16.1 ± 3.0 | 0.015* |
|  | Carbohydrates (g) | 130 | 98.5 ± 17.8 | 97.1 ± 19.1 | 0.768 | 102.8 ± 19.7 | 109.4 ± 22.6 | 0.413 |
| 37–59 |  |  |  |  |  |  |  |  |
|  | Energy (kcal) | 1800 | 1098 ± 163.6 | 1077 ± 171.2 | 0.583 | 1129 ± 169.5 | 1153 ± 186.1 | 0.139 |
|  | Protein (g) | 24 | 8.2 ± 2.3 | 8.5 ± 2.4 | 0.594 | 8.8 ± 2.5 | 18.3 ± 3.0 | 0.016* |
|  | Carbohydrates (g) | 145 | 115.6 ± 20.3 | 116.4 ± 21.5 | 0.799 | 118.9 ± 22.2 | 124.2 ± 24.8 | 0.067 |

**Note:** RDA – Recommended Dietary Allowance; SD – Standard Deviation; kcal – kilocalories; g – grams; \* – statistically significant difference at $p < 0.05$ .

### Micronutrient Intake among Under-Five Children

Table 5 shows that at baseline, there were no significant differences between groups in micronutrient intake across all age categories (p > 0.05). At endline, the intervention significantly increased intake of select micronutrients across all age groups. For children aged 6–11 months, iron intake increased from 8.0 ± 1.3 mg at baseline to 9.5 ± 1.5 mg at endline (p = 0.029), vitamin A from 310 ± 65.2 µg RAE to 380 ± 75.9 µg RAE (p = 0.042), and vitamin C from 35.1 ± 7.9 mg to 46.2 ± 10.3 mg (p = 0.037). In children aged 12– 36 months, significant increases were observed in iron (5.7 ± 1.9 mg to 7.0 ± 1.4 mg, p = 0.038), zinc (2.1 ± 0.9 mg to 2.8 ± 1.0 mg, p = 0.031), and vitamin A (276.1 ± 61.1 µg RAE to 326 ± 65.8 µg RAE, p = 0.044). Among 37–59 months, iron increased from 6.2 ± 1.8 mg to 7.9 ± 2.0 mg (p = 0.029), vitamin A from 292 ± 62.8 µg RAE to 352 ± 68.9 µg RAE (p = 0.037), and vitamin C from 24.1 ± 6.8 mg to 37.6 ± 8.9 mg (p = 0.043).

**Table 5.** Age-specific micronutrient intake among under-five children at baseline and endline.

| Age Group<br>(Months) | Nutrient | RDA | Baseline |  | p-value | Endline |  | p-value |
| --- | --- | --- | --- | --- | --- | --- | --- | --- |
|  |  |  | Control | Intervention |  | Control | Intervention |  |
|  |  |  | (Mean ± SD) | (Mean ± SD) |  | (Mean ± SD) | (Mean ± SD) |  |
| 6–11 |  |  |  |  |  |  |  |  |
|  | Calcium (mg) | 500 | 335 □ ± □ 45.8 | 348 □ ± □ 50.3 | 0.547 | 352 □ ± □ 48.5 | 461 □ ± □ 58.1 | 0.089 |
|  | Iron (mg) | 11 | 8.3 □ ± □ 1.1 | 8.0 □ ± □ 1.3 | 0.692 | 8.7 □ ± □ 1.4 | 9.5 □ ± □ 1.5 | 0.029* |
|  | Zinc (mg) | 3 | 2.5 □ ± □ 0.6 | 2.4 □ ± □ 0.7 | 0.604 | 2.6 □ ± □ 0.8 | 2.8 □ ± □ 0.9 | 0.116 |
|  | Vitamin A (µg RAE) | 400 | 320 □ ± □ 60.7 | 310 □ ± □ 65.2 | 0.838 | 335 □ ± □ 70.4 | 380 □ ± □ 75.9 | 0.042* |
|  | Vitamin C (mg) | 50 | 38.7 □ ± □ 8.2 | 35.1 □ ± □ 7.9 | 0.712 | 41.5 □ ± □ 9.1 | 46.2 □ ± □ 10.3 | 0.037* |
|  | Folate (µg) | 80 | 92 □ ± □ 15.7 | 88 □ ± □ 17.9 | 0.615 | 95 □ ± □ 18.7 | 110 □ ± □ 20.3 | 0.081 |
| 12–36 |  |  |  |  |  |  |  |  |
|  | Calcium (mg) | 700 | 336.0 ± 51.3 | 329.0 ± 53.7 | 0.547 | 319.0 ± 53.7 | 361.0 ± 74.5 | 0.061 |
|  | Iron (mg) | 7 | 5.9 ± 1.3 | 5.7 ± 1.9 | 0.692 | 6.4 ± 1.7 | 7.0 ± 1.4 | 0.038* |
|  | Zinc (mg) | 3 | 2.3 ± 0.5 | 2.1 ± 0.9 | 0.604 | 2.5 ± 0.9 | 2.8 ± 1.0 | 0.031* |
|  | Vitamin A (µg RAE) | 400 | 265.0 ± 54.9 | 276.1 ± 61.1 | 0.838 | 261.0 ± 67.3 | 326.0 ± 65.8 | 0.044* |
|  | Vitamin C (mg) | 40 | 23.4 ± 7.3 | 22.2 ± 6.9 | 0.712 | 25.4 ± 8.1 | 33.9 ± 8.2 | 0.052 |
|  | Folate (µg) | 150 | 93.0 ± 22.7 | 94.0 ± 20.6 | 0.615 | 103.0 ± 33.0 | 129.0 ± 35.6 | 0.071 |
| 37–59 |  |  |  |  |  |  |  |  |
|  | Calcium (mg) | 1000 | 498.0 ± 54.6 | 517.0 ± 59.1 | 0.732 | 516.0 ± 61.8 | 566.0 ± 90.2 | 0.058 |
|  | Iron (mg) | 10 | 6.1 ± 1.6 | 6.2 ± 1.8 | 0.671 | 6.5 ± 1.8 | 7.9 ± 2.0 | 0.029* |
|  | Zinc (mg) | 5 | 3.4 ± 0.7 | 3.6 ± 0.8 | 0.594 | 3.9 ± 0.9 | 4.2 ± 1.1 | 0.062 |
|  | Vitamin A (µg RAE) | 400 | 281.0 ± 61.2 | 292.0 ± 62.8 | 0.742 | 295.0 ± 67.1 | 352.0 ± 68.9 | 0.037* |
|  | Vitamin C (mg) | 45 | 25.3 ± 7.6 | 24.1 ± 6.8 | 0.771 | 27.9 ± 8.2 | 37.6 ± 8.9 | 0.043* |
|  | Folate (µg) | 200 | 96.0 ± 25.3 | 95.0 ± 26.4 | 0.698 | 104.0 ± 29.1 | 142.0 ± 36.8 | 0.064 |

**Note:** RDA – Recommended Dietary Allowance; SD – Standard Deviation; mg – milligrams; μg – micrograms; μg RAE – micrograms of Retinol Activity Equivalents; kcal – kilocalories; g – grams; \* – statistically significant difference at $p < 0.05$ .

## DISCUSSION

Participation in the Positive Deviance Hearth Intervention (PDHI) significantly enhanced core feeding practices among children aged 6–59 months. The intervention resulted in higher meal frequency and increased dietary diversity scores, reflecting improved compliance with recommended complementary feeding protocols and broader inclusion of diverse food groups in children’s diets. These findings are consistent with evidence from community-based PDH programmes, where structured caregiver engagement, experiential learning, and peer-led demonstrations effectively promoted optimal feeding behaviours (21–23). Improvements in responsive feeding and child-specific meal preparation demonstrate the intervention’s capacity to strengthen caregiver feeding strategies, overcoming barriers such as limited nutritional knowledge, culturally entrenched feeding practices, and caregiver time constraints (24–26). The observed improvements are attributable to repeated household visits, practical demonstrations, and participatory problem-solving, which reinforced knowledge translation into practice.

The PDHI also resulted in significant changes in dietary intake patterns, particularly in the consumption of nutrient-dense food groups. Caregivers in the intervention group demonstrated a marked increase in provision of legumes, animal-source foods, and vitamin A–rich and other fruits and vegetables. These shifts suggest that targeted PDH sessions, which emphasize locally available nutrient-rich foods and practical preparation techniques, can significantly diversify children’s diets and improve nutrient adequacy. The absence of significant increases in cereals, roots, tubers, or dairy highlights the program’s focus on addressing dietary gaps most relevant to the local context rather than achieving uniform increases across all food groups. These findings are consistent with prior PDH studies which reported that participatory approaches and peer-led demonstrations enhance caregivers’ capacity to implement nutrition knowledge in a manner tailored to household food availability and cultural preferences (27,28). According to Chek et al. (27), while PDH programs significantly improve dietary diversity and nutrient adequacy among underweight children when effectively implemented and institutionally reinforced, this potent behavioral intervention requires continued supervision, integration with existing nutrition services, and addressing household food insecurity for optimal effect and sustainability to avoid diminished effects after program withdrawal.

Corresponding to improvements in food group consumption, macronutrient intakes, specifically energy and protein, were significantly higher in the intervention group across all age categories. Infants and children aged 6–59 months achieved substantial increases in mean energy and protein consumption compared to controls. These findings were consistent with previous studies’ results (12,16,29). These enhancements likely reflect both caregiver knowledge of portion sizes and meal composition, and the practical guidance provided during PDH sessions, including reinforcement of nutrient-rich combinations and age-appropriate feeding frequency. Children aged 12–36 months and 37–59 months in the intervention group met recommended dietary allowances for energy and protein, while younger infants approached adequacy, indicating age-related differences in feeding practices that may relate to caregiver confidence, complementary feeding initiation, and the physical ability of younger children to consume sufficient quantities. These findings align with prior studies reporting that PDH interventions enhance macronutrient adequacy among undernourished children when education is combined with hands-on, contextually tailored implementation strategies (30,31). However, intake of some macronutrients did not achieve RDA values, and there were no significant differences between intervention and control groups. These results align with other PDH studies (32,33). This might be attributed to differences in locally available and affordable foods and cultural beliefs regarding certain foods. This explains variability in improvements of specific macronutrients across all age groups after six months of PDH intervention. Continued follow-up, individualized counseling, and integration with household food availability remain critical in sustaining gains across all age strata.

Micronutrient intakes also improved significantly in response to the PDHI, with marked increases in iron, vitamin A, vitamin C, and zinc among children in relevant age groups. By endline, many children in the intervention arm achieved recommended dietary allowances, reflecting the effectiveness of participatory nutrition education in promoting consumption of locally available, nutrient-dense foods. The increased intake of legumes, animal-source foods, and vitamin A–rich vegetables likely contributed directly to improved micronutrient profiles, as observed in previous PDH studies across diverse settings (34–36). These studies reinforce the premise that experiential learning, peer-led counseling, and practical demonstrations not only improve food diversity but also enhance the adequacy of critical micronutrients linked to growth and immunity. Nonetheless, evidence suggests that sustained engagement and program reinforcement are critical to maintaining these gains, as improvements in micronutrient intake may diminish without continuous monitoring, institutional anchoring, and integration with broader nutrition or agricultural interventions (13,36,37). Similarly, longitudinal evidence indicates that micronutrient gains are not uniformly sustained. Csoelle et al. (37) reported declines in iron and zinc intake within one year of program withdrawal where follow-up was absent. Putri et al. (13) observed durable improvements only in settings where PDH counseling was integrated into routine community nutrition services. These findings suggest that PDHI effectiveness on micronutrient adequacy is contingent on local food availability, dietary bioavailability, and institutional continuity. The combination of targeted counseling, hands-on demonstrations, and peer support likely accounts for the magnitude of observed changes, particularly for nutrient-dense foods that were previously under-consumed. These findings provide strong evidence that community-based, positive deviance approaches can produce measurable nutritional benefits among malnourished children, and emphasize the importance of integrating such programs within ongoing public health, nutrition, and community development initiatives to ensure sustainability and long-term impact.

### Study Strengths and Limitations

This study has various strengths. The study adopted validated tools and structured measurement of feeding indicators. The hearth and peer-led sessions strengthened caregiver engagement and cultural relevance. The implementation of intervention and longitudinal follow-up up to six months enabled assessment of both short- and mid-term outcomes of the PDH intervention. The intervention was implemented in a high-burden rural setting County, hence strengthening ecological validity. Additionally, the study findings demonstrated feasibility and scalability of PDH intervention through integration to existing community health approaches and routine community nutrition approaches strengthening its programmatic relevance. However, the study was limited to one sub county in the county of study influencing generalizability of findings to different contexts and regions. The study was conducted for six months hence potentially not fully capturing long-term sustainability. Additionally, the reliance of caregiver-reported practices for some indicators introduces potential recall and social desirability bias. However, the study ensured quality data were collected.

## Conclusion

This study demonstrates that the Positive Deviance Hearth intervention effectively improves child feeding practices in a high-burden rural setting. Significant gains were observed in meal frequency, dietary diversity, and responsive feeding, indicating successful adoption of recommended behaviors by caregivers. The community-based, peer-led approach facilitated practical learning and enhanced acceptability, supporting its relevance for low-resource contexts. Improvements in feeding practices were accompanied by increased consumption of nutrient-dense foods, suggesting potential for sustained dietary adequacy. However, persistent structural barriers, including food insecurity and economic constraints, may limit long-term adherence and scalability. The findings underscore the importance of integrating behavior-focused nutrition interventions within existing community health systems. Future programs should incorporate strategies addressing household-level constraints to strengthen sustainability. Additionally, PDH programs should be embedded in community health systems with scalable caregiver training, peer-led demonstrations, and continuous mentorship to sustain improved feeding and hygiene practices long-term.

## Data Availability

All data produced in the present study are available upon reasonable request to the authors

